# Echo-planar imaging-based time-of-flight imaging using a modified interleaved flyback trajectory

**DOI:** 10.1101/2025.10.29.25339048

**Authors:** Simon Blömer, Tony Stöcker, Rüdiger Stirnberg

## Abstract

**Purpose:** A modified interleaved flyback (miFB) approach is introduced, designed to mitigate flow artifacts caused by alternating readout polarities in Echo Planar Imaging (EPI), while preserving acquisition efficiency.

**Methods:** We propose reconstructing odd and even echoes of 3D-EPI separately. To this end, the respective missing lines are acquired in interleaved shots with inverted polarity and an additional precursor gradient. Thereby, high scan efficiency is maintained compared to unsampled flyback gradients. Our miFB approach is additionally combined with gradient moment smoothing and compared to the interleaved dual-echo with acceleration (IDEA) method in phantom and in vivo scans at 7 Tesla.

**Results:** Our results demonstrate a significant reduction in ghosting and signal dropout using the miFB approach, yielding comparable image quality to non-EPI acquisitions while reducing acquisition time by approximately half.

**Conclusion:** The miFB approach offers a substantial reduction in flow artifacts, allowing for decreased acquisition times in TOF-MRA.

## 1. Introduction

Intracranial Time-of-flight (TOF)-MRA is a challenging procedure due to its long scanning times. However, Compressed Sensing (CS) has been shown to reduce acquisition times while maintaining comparable diagnostic performance (1,2). EPI can substantially accelerate acquisition times by acquiring multiple lines of k-space during each TR (3). Early applications of 3D segmented EPI have demonstrated its usefulness for whole heart MRA (4,5). The combination of EPI with controlled aliasing in parallel imaging (CAIPIRINHA) (6) enables higher acceleration factors, leading to more efficient sampling and reduced acquisition times (7–9). Skipped-CAIPI sampling further enhances flexibility by allowing for optimal adjustment of sequence parameters, such as EPI factor and phase encode bandwidth, combined with a suitable CAIPI pattern (10). Although EPI enables very efficient and fast sampling of k-space, it comes at the cost of increased vulnerability to flow artifacts. Flow during application of encoding gradients in multi-shot EPI leads to phase errors, causing signal loss and ghosting (3,11–14). Two main artifact sources have been identified: flow along the phase encoding axis (y, w.l.o.g) (12,15,16) and flow along the frequency encoding axes (x, w.l.o.g) (11,12,15). To mitigate the effects of flow along the y-axis, compensation strategies such as centric reordering or partial Fourier sampling (17,18) and gradient moment smoothing have been proposed (16). However, flow along the frequency encoding axes poses a distinct challenge, as phase errors accumulate differently during odd and even readouts, leading to ghosting artifacts. The flyback approach mitigates this issue but reduces scanning efficiency, since no echo signals are acquired during flyback gradients (11). Particularly when running an EPI sequence close to hardware limitations, e.g. for high resolutions, flyback gradients and readout gradients have comparable durations such that half of the potential signal acquisition window remains unused. To overcome this limitation, we propose a modified interleaved flyback trajectory (miFB) that uses two regular EPI acquisitions but reconstructs odd and even echoes separately, allowing for ghosting reduction while preserving efficiency through optional averaging (19). Notably, our miFB approach shares similarities with the previously introduced IDEA approach (20).

## 2. Method

### 2.1 Mitigation of flow artifacts in EPI

The phase evolution of a moving spin during a time-dependent gradient waveform 𝐺(𝑡) can be expressed using the Taylor series expansion as follows:

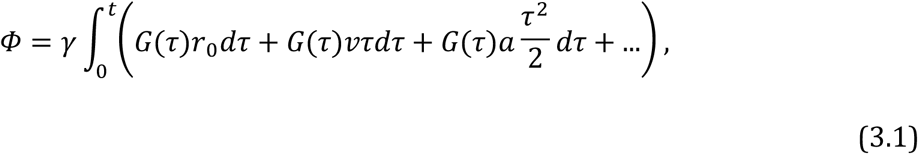

where 𝛾 is the gyromagnetic ratio and 𝑟_0_,𝑣, and 𝑎 are the spatial position, velocity, and acceleration of the moving spin, respectively, along the direction *G*. Oscillating readout gradients in the x-direction used in EPI toggle between positive and negative gradient amplitudes to achieve fast encoding of k-space. This toggling, however, leads to ghosting artifacts in images, as shown by Duerk and Simonetti (11) and Butts and Riederer (12), if there is non-zero velocity along x. Phase, proportional to the velocity, *v*, accumulates during all odd echoes, resulting in a phase offset at the midpoint of these echoes. Using a precursor gradient lobe, the phase offset can be shifted between even and odd echoes (3). Given that equal amplitude (with alternating sign) and durations are applied for all readout gradients, the accumulated phase is subsequently reversed during even echoes, refocusing the phase offset near the center of the gradients. The phase difference between odd and even echoes at *k_x_*=0 can be determined by comparing their first-order gradient moments (12). Using the properties of the readout gradients, such as rise time 𝑟 and amplitude 𝐺_𝑚𝑎𝑥_, and the echospacing, ESP, we can express the phase difference as:

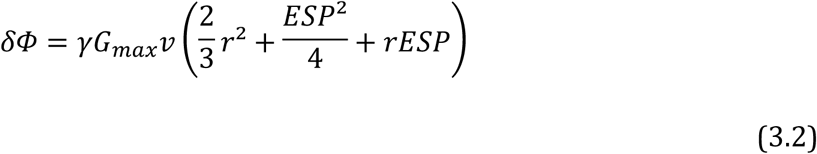

These phase discontinuities between adjacent lines or echo segments for single-shot and interleaved multi shot EPI, cause significant ghosting artifacts scaling with the velocity along the frequency-encode axis.

### Modified interleaved flyback (miFB)

The acquisition of two temporary k-spaces, denoted as k_initial_ and k_repeated_, is achieved by performing each segmented EPI shot twice in an interleaved fashion. With the initial shots, a full k_x_,k_y_ plane in k_initial_ is acquired using the regular segmented EPI sampling scheme, as illustrated in the sequence diagram in Figure 1A. With the repeated shots, the same k_x_,k_y_ plane is acquired in k_repeated_ with two key modifications:

1. All readout polarities are inverted.
2. A precursor gradient lobe (Figure 1A, black dotted) is added before the first readout, introducing an unsampled echo that effectively swaps odd and even echoes.

**Figure 1:**
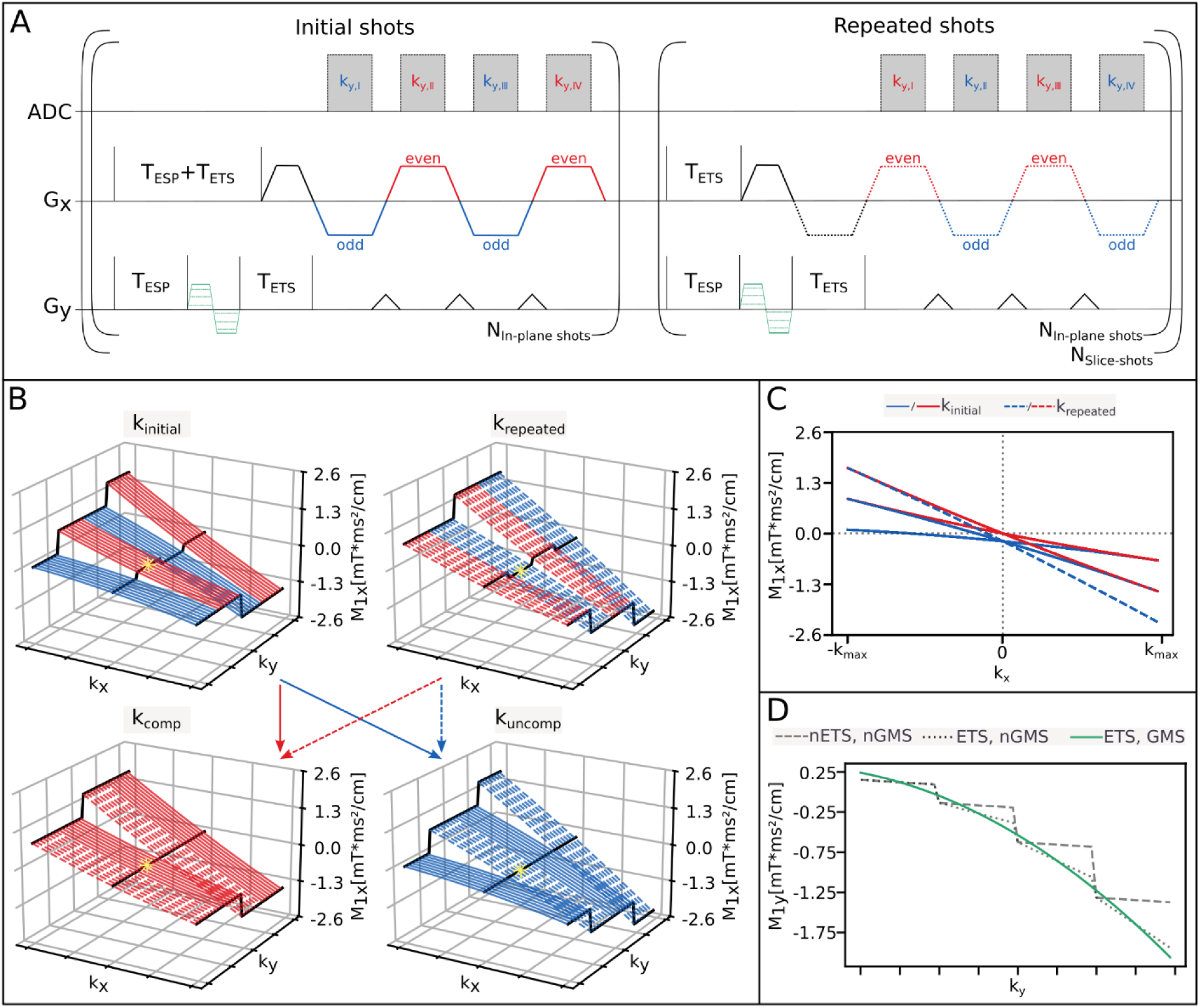
(A) Sequence diagram of the miFB approach, illustrating the encoding gradients for the x- and y-axes (Gx, Gy). The two interleaved acquisitions of the same shot are shown, with an unsampled precursor gradient lobe added in the repeated shots and an additional echospacing (ESP) applied in the initial shots. The additional GMS gradient is shown in green. (B) Simulated first-order moments from flow along the x-axis for the acquired kinitial, krepeated, and shuffled kcomp and kuncomp k-spaces. An asterisk indicates the center of k-space for acquisitions using 6/8 phase partial Fourier sampling. (C) Combined projection of the simulated moments along the y-axis. (D) Phase evolution for flow along the y-axis, demonstrating the effects of different combinations of echo train shifting (ETS) and gradient moment smoothing (GMS). Note that on the y-axis, the duration of TETS as well as the GMS gradient (green) is varied for each shot (A). The quadratic steps apparent without ETS and GMS (nETS, nGMS) are reduced to steps of equal height when ETS is activated, but GMS is not (ETS, nGMS). The use of both ETS and GMS results in a smooth quadratic M1 curve (ETS, GMS).

From the temporary k-spaces, all signals acquired with positive gradient polarity are reordered to one of two final flyback k-spaces, k_comp_ and k_uncomp_, and signals acquired with negative gradient polarity to the other. Readout gradients of k_uncomp_ served as flyback gradients for k_comp_. In addition to a consistent frequency-encode polarity, the unsampled precursor gradient lobe assures that all echoes in the interleaved flyback k-spaces are acquired with a consistent moment at k_x_=0.

An additional dead-time of one ESP before the prephasing gradient of all initial shots in combination with consistent echo time shifting (21) ensures the same linear TE increase along k_y_ at k_x_=0. Furthermore, due to the consistent frequency-encode polarity, the final 3D images miFBc and miFBuc reconstructed from k_comp_ and k_uncomp_, respectively, do not suffer from off-resonance-induced segmentation artifacts (22). Note that both images have the same TE.

### Interleaved dual-echo with acceleration EPI (IDEA-EPI)

IDEA-EPI (20) was originally introduced to reduce artifacts from gradient imperfections and off-resonance effects that cause inconsistencies between odd and even k-space lines. Similar to our miFB approach, two k-spaces are acquired and reconstructed separately. For IDEA, instead of acquiring each shot twice in two interleaves, every other phase encoding blip is removed, yielding two k-spaces with different TEs. This results in twice as many frequency-encoding lobes per shot, doubling the echo train length, echo spacing, and phase evolution. Initially, it was proposed to use parallel acceleration to skip half of the k-space lines (20). However, given that we already employ parallel acceleration, this approach may not be feasible. Therefore, we suggest doubling the segmentation factor to acquire all skipped lines in subsequent shots.

### Gradient moment smoothing (GMS)

First order moments from flow along the y-axis increase quadratically during phase encoding blips resulting in a large phase offset at the k-space center. Interleaved multi-shot EPI increases the complexity of the phase evolutions, introducing step-like phase variations between echo segments (12). Using a “sliding-readout-window” (23) approach also called “echo-time-shifting” (21) reduces phase variations, although – due to the quadratic increase in height – steps of linear height remain (16), which cause residual ghosting. First order GMS can be employed, assuring that the first order moments of all phase encoding blips lie on a smooth quadratic curve without steps (no residual linear M_1,y_ jumps according to Fig. 1D). Instead of attempting to null the resulting phase offset at k_y_=0 through time-consuming bipolar gradients, as described in (16), we propose to use GMS only to eliminate the residual linear phase discontinuities as described. However, throughout this work, the overall phase offset is kept as small as possible through partial Fourier acquisition along k_y_ (15).

### 2.2 Simulations

Our simulation was limited to first-order moments, with the assumption that higher-order effects are negligible. The first-order moments of flow along the x-axis were computed using formula 3.1 and the specific gradient waveforms and sequence timings of the unmodified EPI sequence and the miFB sequence. Assuming rectangular gradient (w.l.o.g.), the first-order moment from the y-blip of the *k*th gradient echo of the *n*th shot is given by the sum of the moments resulting from the PE-prephaser 𝑀_1𝑦_(1,𝑛), the current blip and all previous blips:

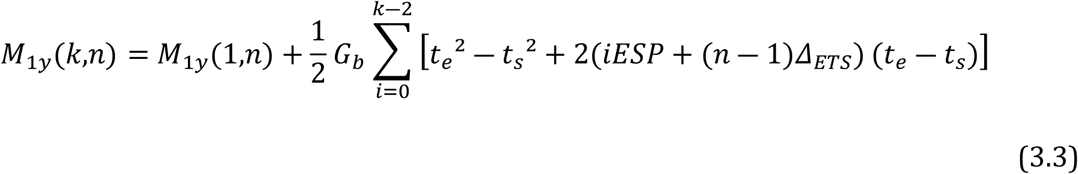

(for 2 ≤ k ≤ ETL and 1 ≤ n ≤ N), where 𝐺_𝑏_ is the amplitude of the y-blips, 𝑡_𝑠_ and 𝑡_𝑒_ are the start and end times of the first blip of the first shot and 𝛥_𝐸𝑇𝑆_is the TE increment due to echo time shifting (16). The simulation parameters were chosen to match those of the 0.6 mm EPI sequences in the in vivo protocol, with a reduced matrix size of 64x64. The amplitude of the readout and phase encoding gradients were set at 34.5 and 34 mT/m, respectively. M_1_ from flow along the x and y-axis were simulated separately and are shown in Figure 1B, C for x-flow and 1D for y-flow.

### 3.4 Experiments

Flow phantom (Figure 2) and in vivo measurements were performed on a MAGNETOM 7T Plus scanner (Siemens Healthineers, Erlangen, Germany) using a 32-channel head coil for receive, and a volume coil for transmit (Nova Medical, Wilmington, MA). Ten healthy volunteers were scanned after providing written informed consent. Three time-of-flight (TOF) sequences with in-plane isotropic resolutions of 0.6 mm, 0.5 mm, and 0.4 mm and slice resolution of 0.8 mm were acquired. A custom skipped-CAIPI 3D-EPI sequence (hereafter referred to as “EPI”) was used for shot-selective CAIPI sampling with EF=4 and a 1x3_z1_ CAIPI pattern. Besides regular EPI, optional miFB with or without GMS and IDEA sampling were implemented. Two additional sequences were employed as references: a modified vendor-provided standard GRE sequence using the same CAIPI pattern and image reconstruction as EPI (referred to as “GRE”), and a vendor-provided GRE sequence optimized for TOF imaging (referred to as “Vendor”). All three resolutions were acquired using EPI with miFB and GMS activated (miFB+GMS), as well as the IDEA approach. At 0.6 mm resolution, two additional EPI scans were performed: one without miFB but with activated GMS (EPI+GMS), and another scan with miFB and deactivated GMS (miFB). The x, y FOV was 200 × 200 mm for all measurements, with the x-axis oriented in the left-right direction and the y-axis in the anteroposterior direction. With EPI, a slab-selective binomial-121 water excitation pulse with a nominal flip angle of 24 degrees and a Sinc time-bandwidth product 6 was used while GRE used a regular Sinc pulse with a lower time-bandwidth product of 5.12 but the same nominal flip angle. The Vendor TOF sequence used a TONE excitation pulse with a TONE ramp of 70% and flip angle of 20 degrees. Flow direction was set to head-feet. The repetition time (TR) was optimized for each sequence: EPI and GRE used a TR of 18 ms, whereas Vendor required an increased TR of 30 ms due to SAR limitations. At a resolution of 0.4 mm, the TR had to be increased for all sequences, and the flip angle was adjusted accordingly. The slice thickness was set to 0.8 mm for all three resolutions measuring 87 slices per slab for EPI, and 88 slices per slab for GRE and Vendor. Flow compensation was activated for GRE along x and Vendor along all directions. Further sequence parameters can be found in Table 1. In addition to measurements with in-plane resolutions of 0.6 mm, 0.5 mm, and 0.4 mm, miFB+GMS was acquired at 0.8 mm with comparable sequence parameters to the ones described above. For anatomical reference, a 3D-T_1_-weighted magnetization prepared rapid gradient echo (MP2RAGE)(24) sequence was acquired at 0.6 mm isotropic resolution. All images were reconstructed using vendor implementations of GRAPPA (25), with additional 2D CAIPIRINHA (26) (IcePAT) support for EPI and GRE. The vendor’s default phase correction and ramp sampling were utilized.

**Figure 2:**
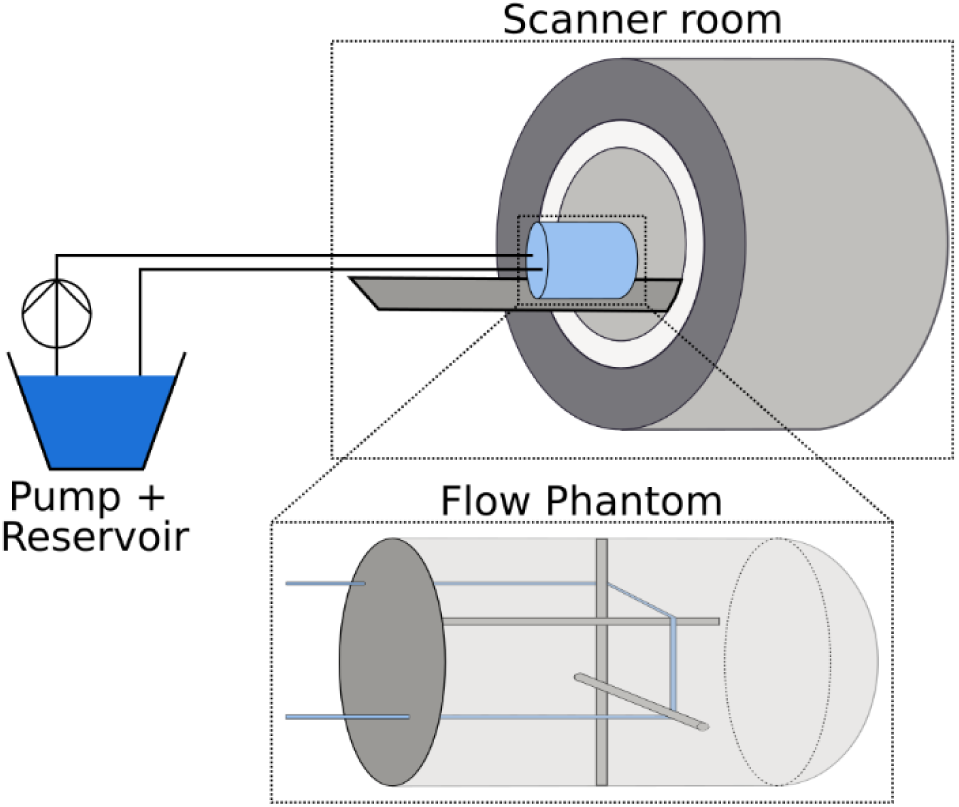
Schematics of the flow phantom setup used in the phantom experiment. A peristaltic pump (Thermo Fisher Scientific FH100), located outside the scanner room, circulated distilled water through a 1.5 mm inner diameter PVC tube embedded in a cylindrical phantom filled with a PVP–agarose–NaCl mixture. The tube was arranged around internal supports to produce simultaneous flow along three orthogonal directions, as illustrated. The pump flow rate was calibrated to a constant velocity of approximately 35 cm/s. Phantom measurements were acquired in two configurations: (1) with the flow aligned along the scanner’s principal encoding axes, and (2) with the phantom rotated to create oblique flow relative to the frequency and phase encoding directions, enhancing visibility of y-flow-induced artifacts.

For miFB and IDEA, the two k-spaces (k_comp_ and k_uncomp_) were reconstructed separately into two 3D images. All 0.6mm scans and the protocol described above were repeated for the phantom measurements and the two tubing orientations at a constant velocity of approximately 35 cm/s. In vivo images were preprocessed using N4ITK bias-field (27) and non-local means (28) from the Advanced Normalization Tools (ANTs) framework in Python (29). Vessels were segmented using the pretrained deep-learning model “predict” from the Python-based VesselBoost package (30) with preprocessing disabled.

### 3.5 Evaluation

To compare the effect of the miFB and IDEA sampling, the segmented images were evaluated at the M1 segment of the middle cerebral artery (MCA) since largest flow artifacts were observed in the EPI images without flyback here. Vessel diameters were calculated to evaluate the artifact suppression capabilities of the sequences. An adaptation of the processing pipeline described by Avadiappan et al. (30) in Python was used (Figure 3). To account for minor segmentation differences between TOF images, 2D vessel diameter map were divided into 90 vertical bins of equal size. Gold-standard-to-difference plots were created comparing the difference between the mean diameter of each bin of the EPI and GRE sequences and the gold standard Vendor TOF on the y-axis to the mean diameter of each bin in the Vendor TOF on the x-axis. The two flyback images were analyzed individually.

**Figure 3:**
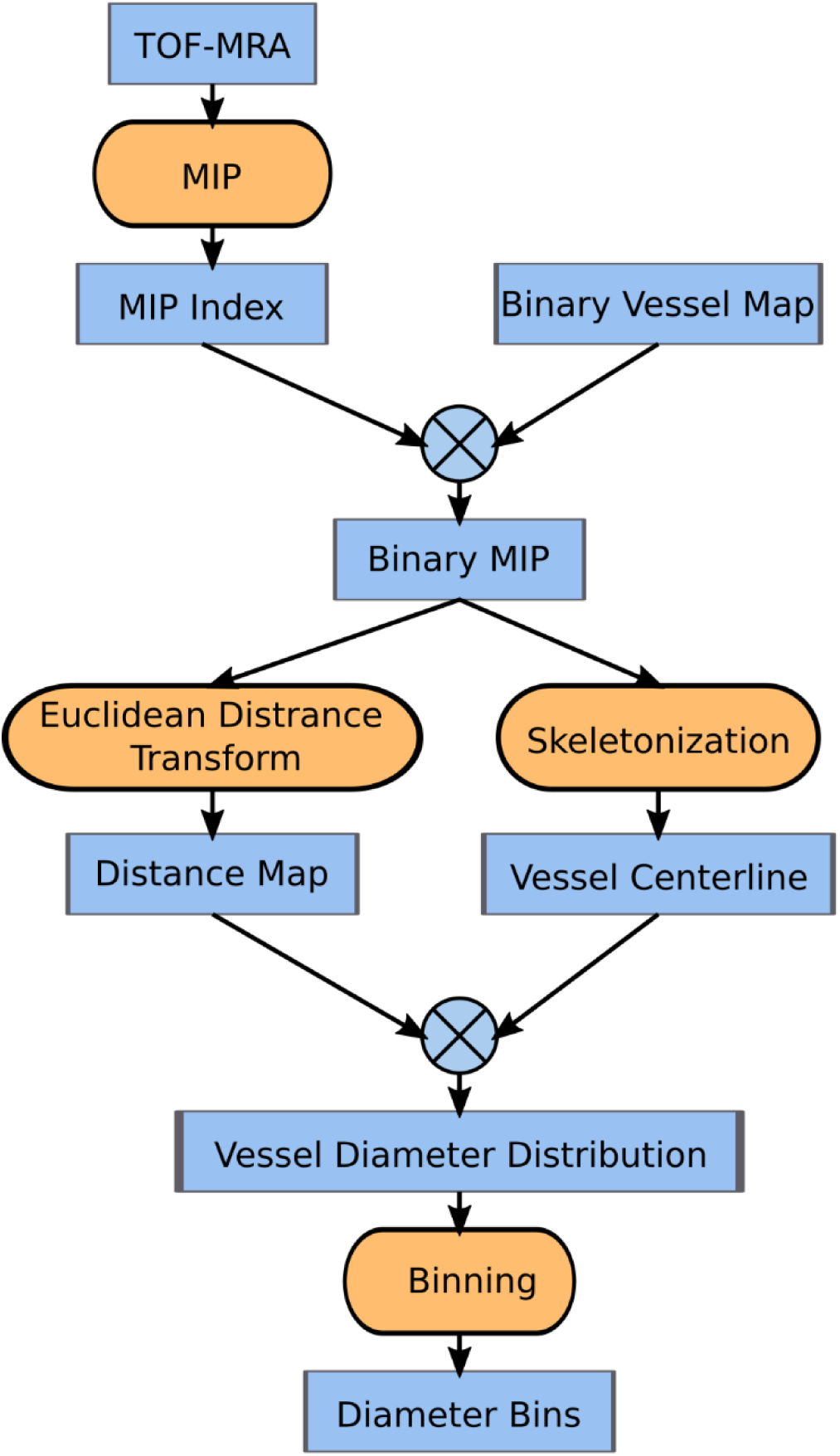
Evaluation pipeline for the MCA diameter computation. Maximum intensity projections (MIPs) are generated along the head–feet direction. The indices of maximum intensity pixels along this axis are recorded and applied to the corresponding segmented vessel images (from preprocessing) to obtain MIPs of the segmentations. A 2D Euclidean distance transform (EDT) is applied to these segmented MIPs. Vessel centerlines are extracted by skeletonizing the binary segmentation maps. Diameters are computed by sampling the EDT map at the skeleton positions. To account for minor segmentation differences between TOF images, the resulting 2D vessel diameter map is divided into 90 bins of equal size along the left-right direction. For each section, the mean diameter is calculated over all non-zero pixels.

To evaluate the effect of the additional GMS gradient, a similar process was performed manually using the Vascular Modeling Toolkit (VMTK) (31) of 3DSlicer (https://www.slicer.org/) v5.8.1(32–35) evaluating the circular equivalent (CE) diameter and cross-section area of the ACA in each scan. For each TOF-image, the median over all these values was calculated. Gold-standard-to-difference plots were created as described above.

To quantitatively assess the difference between the compensated and uncompensated miFB+GMS images, we computed the Structural Similarity Index Measure (SSIM) across four resolutions and visualized the results. The compensated and uncompensated images are shown alongside their respective mean images for one exemplary subject.

## 3. Results

### 3.1 Simulations

Figure 1B, C shows the simulated first order moments M_1,x_ from flow along the x-direction for the miFB approach. Both temporary k-spaces show oscillating phase behaviors between odd and even echo segments at k_x_=0. All odd echoes in k_initial_ acquire a negative phase offset, that is reversed by even echoes (red). Adding a precursor gradient lobe shifts the phase offset to even echoes (red) in k_repeated_. The increased time between prephaser and first readout leads to a larger M_1_ for all echoes in k_repeated_ compared to k_initial_. In fact, since the additional time is exactly one ESP, all n-echoes in k_repeated_ show the same phase evolution as all (n+1)-echoes in k_initial_. This can be seen in Figure 1C, where only the moments of the first echo in k_initial_ and last echo in k_repeated_ are not overlayed by another echo of k_repeated_ and k_initial_, respectively. Adding the shot specific ETS delay and additional ESP of the initial shots before the prephasing gradient assures that all initial and repeated shots share the same reference time point. After reordering, both k_comp_ and k_uncomp_ show a smooth moment evolution at k_x_=0, that is zero for k_comp_ with a negative offset for k_uncomp_.

For k_x_≠0 discontinuities remain, that have the same amplitude and k_y_ spacing for both k-spaces, though appear at different positions and frequency along k_y_. In k_comp_ only one discontinuity can be observed at k_y_=0, while two discontinuities can be observed at k_y_=1/3 and k_y_=2/3 for k_uncomp_. Note that, due to 6/8 phase partial Fourier sampling in the actual experiments, k_y_=0 in the simulations is shifted with respect to the actual k_y_=0 in the experiments (the simulated k_y_ range covers 6/8 of the experimental k_y_ range).

In Figure 1D simulation results for M_1,y_ from flow along the y-axis are shown. EF-1 steps can be observed, that increase quadratically for the case without ETS and GMS (dashed) and are of equal height for the case without GMS but ETS activated (dotted). Using both ETS and GMS leads to a smooth quadratic M_1_ curve (green). Note, since the additional time delay introduced by ETS between the GMS gradient, respectively prephasing gradient and first PE blip is close to one ESP for the last shot, M_1_ at the end of the echo-train is significantly larger for both cases with ETS enabled compared to the case without.

### 3.2 Phantom results

Phantom results are shown in Figure 4 as maximum intensity projections (MIP) along the z-axis. For all flyback scans, images reconstructed from compensated (k_comp_, red) and uncompensated (k_uncomp_, blue) k-spaces are overlaid for comparison. With flow parallel to the y-axis (Fig. 3A, ∼0° tubing angulation), no significant differences are observed among all compared sequences, as ghosting artifacts from flow along y propagate parallel to the y-axis overlapping with signal from the tubing. However, for tubing oriented parallel to the x-axis, substantial ghosting artifacts and signal loss becomes apparent in EPI+GMS (green arrow). In contrast, all compensated flyback images (miFBc+GMS, miFBc and IDEAc) exhibit comparable results to those obtained with the GRE and Vendor protocols. Notably, significant signal loss is evident with flow along x in all uncompensated flyback images (miFBuc+GMS, miFBuc and IDEAuc) (orange arrow). Flow angulated at approximately 45 degrees to both x- and y-axis (Fig. 3B) yields similar artifact-free results for miFBc+GMS, as well as for the GRE and Vendor protocol, comparable to the non-angulated case. In all other EPI measurements, GMS was deactivated, resulting in ghost artifacts observable in all images (IDEAc, miFB_c,uc_, and EPI+GMS) when flow is directed at 45 degrees to both x and y-axis (yellow arrow). For IDEA, spacing between ghosts is smaller compared to the other two sequences, since for IDEA the EF and hence the number of blips and k-space steps is reduced by a factor of two. Like the 0-degree case, signal loss is evident in all uncompensated flyback images (miFBuc+GMS, miFBuc and IDEAuc).

**Figure 4:**
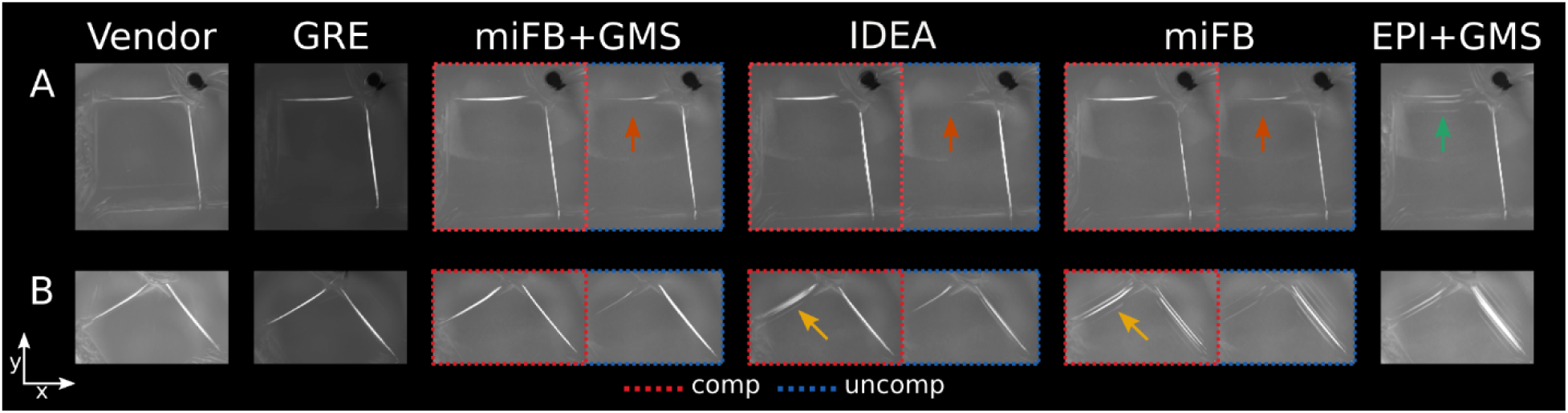
Zoomed-in sections of the maximum intensity projections along the z-axis are shown for two flow phantom orientations: flow approximately aligned with the scanner’s encoding axes, and flow at an oblique angle (∼45 degrees) to the encoding axes, highlighting y-flow induced artifacts. The results are compared across different sequences, including miFB with GMS (miFB+GMS) and without (miFB), as well as IDEA, GRE, Vendor, and regular EPI GMS (EPI+GMS). For the miFB and IDEA sequences, the two flyback images (compensated, red; and uncompensated, blue) are overlaid for visual comparison.

### 3.3 In vivo results

Figure 5 shows corresponding results of in vivo measurements for one volunteer at 0.6 mm, displayed as maximum intensity projections (MIPs) along the z-axis. miFB together with GMS (miFBc+GMS) and Vendor exhibit similar artifact-free results. However, significant ghosting artifacts are observed for EPI without the miFB approach (EPI+GMS) at the level of M1 in the MCA, marked by a yellow arrow, where flow is primarily directed along the x-axis. IDEAc shows widening of the MCA and posterior cerebral artery (PCA), with the right superior cerebral artery being overlaid by the right PCA (marked by an orange arrow). This might be the result of the reduced distance between ghosts resulting from EF of two and the spacing between ghosts being smaller than one pixel. Like the phantom results, significant signal loss is observed in vessels oriented primarily along x (e.g. MCA), for all uncompensated flyback images (miFBuc+GMS, miFBuc, and IDEAuc). In contrast, less signal loss is visible in the PCA, as it is primarily directed along the y-axis. For GRE, reduced signal is found at all resolutions for vessels close to the boundaries of the FoV along the non-blipped phase encoding axis (z, w.l.o.g.), such as the PCA. For sequences that do not employ flow compensation along the z-direction (all except Vendor), signal loss can be observed for the internal carotid artery (ICA). This flow-induced signal loss gets more pronounced at increasing resolution. miFB without GMS (miFBc) leads to ghosting for vessels that are oriented along both the readout and y-direction, e.g. right MCA as indicated by the yellow arrow. Note, that for the left MCA, oriented almost exclusively along the x-axis, no ghosting is visible. For vessels that are aligned primarily along the y-axis, ghosts overlap with the vessel and are not visible. Figure 6 shows the results at resolutions of 0.5, and 0.4 mm. At a resolution of 0.5 mm, similar image quality is observed for all measured sequences compared to the 0.6 mm measurements. However, at 0.4 mm, significant signal loss is evident for miFB(c,uc)+GMS in vessels oriented mainly along the y-axis (e.g. PCA and ACA). This is not the case for IDEAc, possibly due to the reduced number of PE-blips.

**Figure 5:**
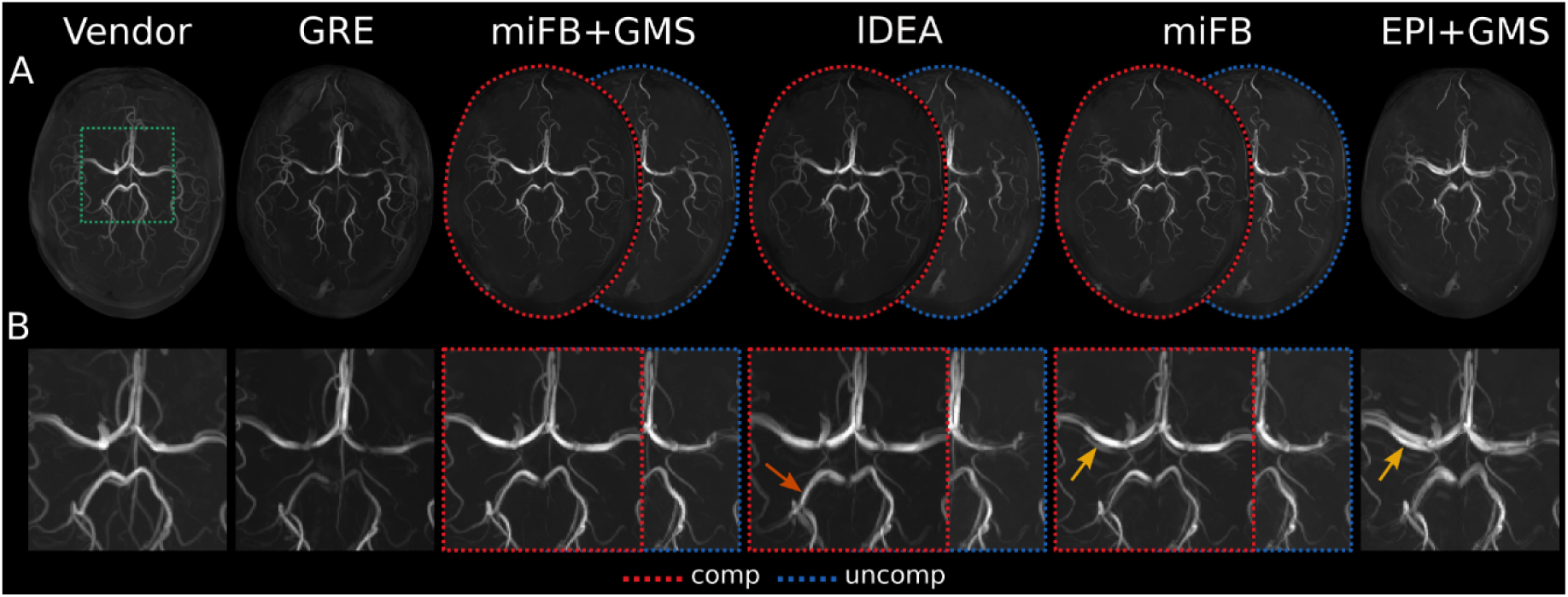
Results of in vivo measurements for one volunteer at 0.6 mm, displayed as MIPs along the z-axis. miFB with GMS (miFB+GMS) and without (miFB) is compared to IDEA, GRE, Vendor and regular EPI with GMS (EPI+GMS). The two flyback images (compensated, red; and uncompensated, blue) are overlaid for comparison. Whole brain MIPs (A) and a zoomed section (B), indicated by a green box in (A), are shown for better visualization. Ghosting artifacts are visible for EPI+GMS and miFB, as indicated by the yellow arrow. Additionally, widening of the posterior cerebral artery (PCA) is observed for IDEA, marked by an orange arrow.

**Figure 6:**
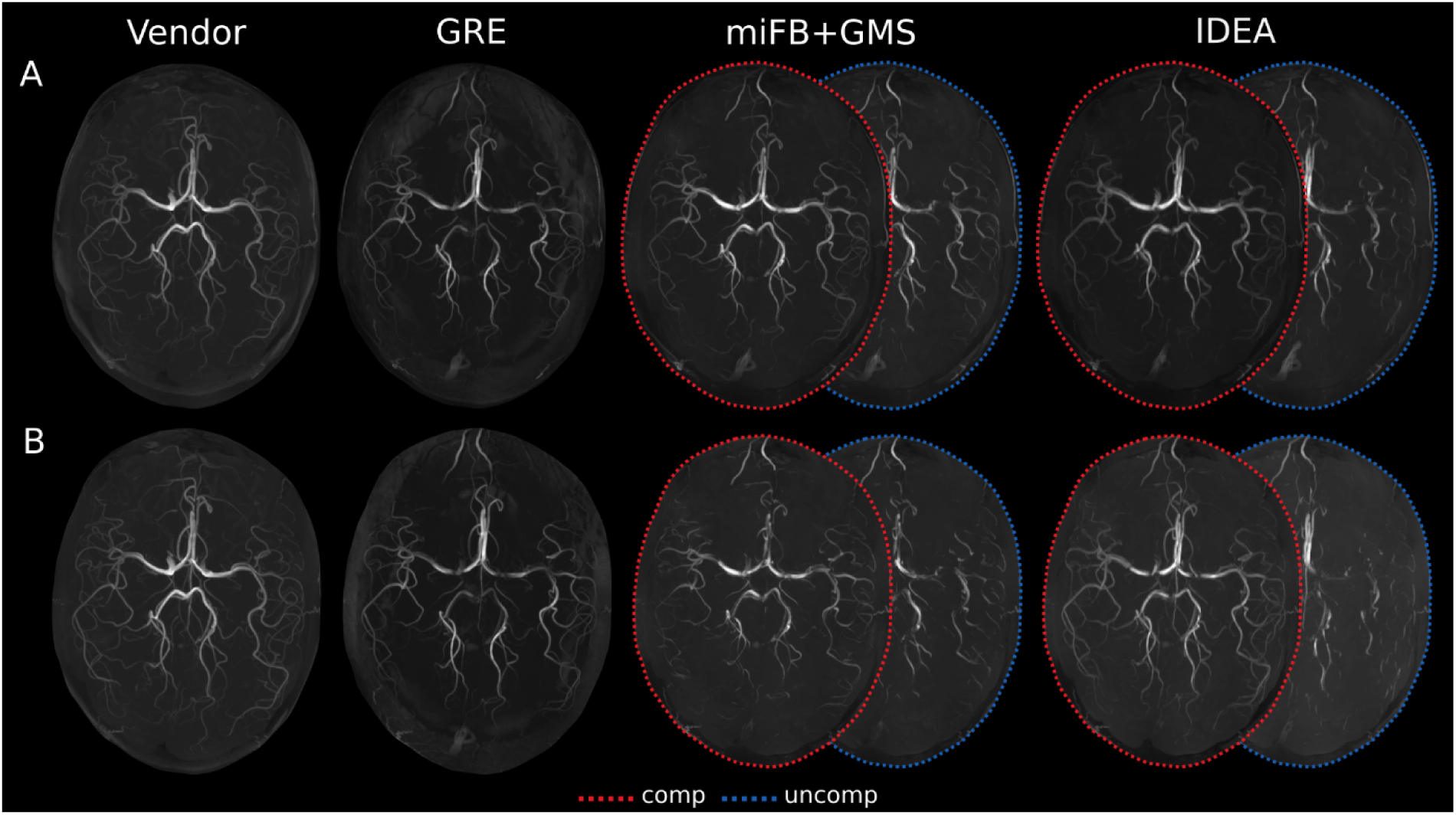
Results of the in vivo measurements for 0.5 mm in A and 0.4 mm in B. The two flyback trajectories miFB with GMS and IDEA (compensated, red; and uncompensated, blue) are compared to GRE and Vendor. Significant signal loss can be observed in uncompensated images. At a resolution of 0.5 mm, similar image quality is observed for all measured sequences. Larger y-flow induced artifacts appear in the compensated miFB+GMS image compared to IDEA for 0.4 mm.

Similar to IDEAuc, vessels oriented along the x-axis show significant flow artifacts whereas vessels oriented along the y-direction do not. Increasing the resolution leads to more signal loss in uncompensated flyback images for vessels oriented along the x-axis, whereas GRE and Vendor exhibit no decrease in image quality with increasing resolution. Compared to all other sequences, Vendor exhibits enhanced visibility of small vessels at all resolutions.

The relationship between resolution and mean SSIM, comparing compensated to uncompensated miFB+GMS images, is illustrated in Figure 7. A non-linear increase in SSIM is observed as resolution decreases, converging toward a value near 1. Mean images from a representative volunteer show a substantial reduction in flow artifacts with increasing voxel size, resulting in near-identical image quality between the compensated and mean images at in-plane isotropic resolutions of 0.6 mm and 0.8 mm.

**Figure 7:**
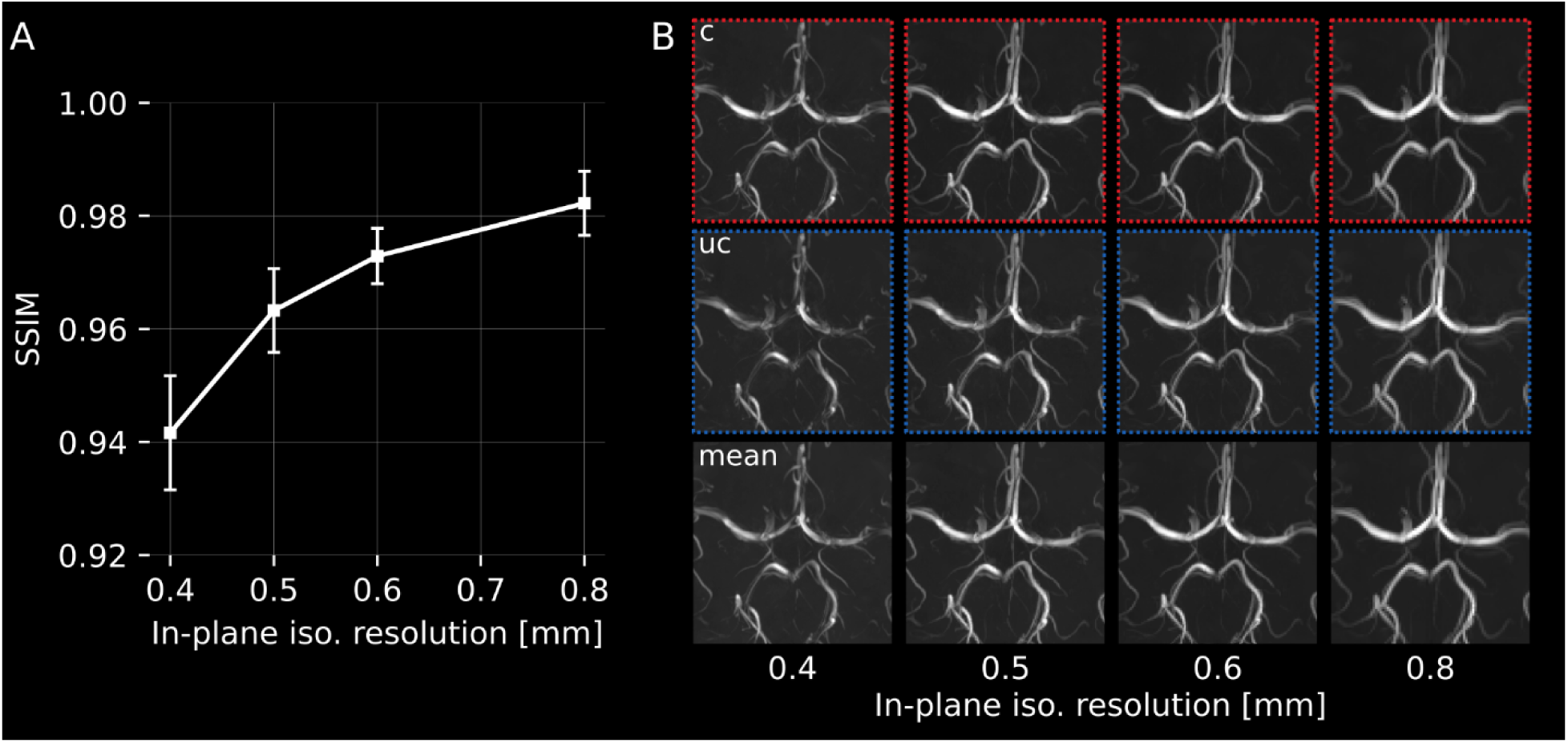
(A) Group mean and SD of the SSIM between the two miFB+GMS images as a function of the in-plane isotropic resolution comparing compensated to uncompensated images. SSIM was evaluated at four resolutions (0.4 mm, 0.5 mm, 0.6 mm, and 0.8 mm) across all volunteers. A non-linear increase in SSIM with increasing voxel size is observed converging toward a value near 1. (B) Zoomed-in sections from the two miFB+GMS images acquired at the same four resolutions. The MIP from the compensated image (red) and the uncompensated image (blue) are shown for each resolution. The MIPs of the corresponding mean images are displayed in the bottom row. As voxel size increases, flow-related artifacts in the uncompensated images are visibly reduced, resulting in improved quality of the mean images.

Gold-standard-to-difference plots for the estimated MCA diameter are presented in Figure 8, with Vendor considered as the gold standard. Mean and SD of the differences are shown. Similar difference distributions were found for all flyback images at 0.6 mm resolution (A), with the smallest differences observed for miFBc+GMS. Ghosting was observed for EPI+GMS, leading to larger deviations in vessel diameters. A linear dependency of the difference to Vendor was observed for all images, including GRE, as indicated by the yellow line, stemming from segments where vessel information is fully destroyed by flow artifacts, resulting in a vessel diameter of zero. The largest number of missing vessels at 0.6 mm resolution was found for GRE, miFBuc, IDEAuc, and EPI+GMS. At 0.5 mm resolution (B), similar differences were found for GRE, miFB, and IDEA compared to the 0.6 mm case.

**Figure 8:**
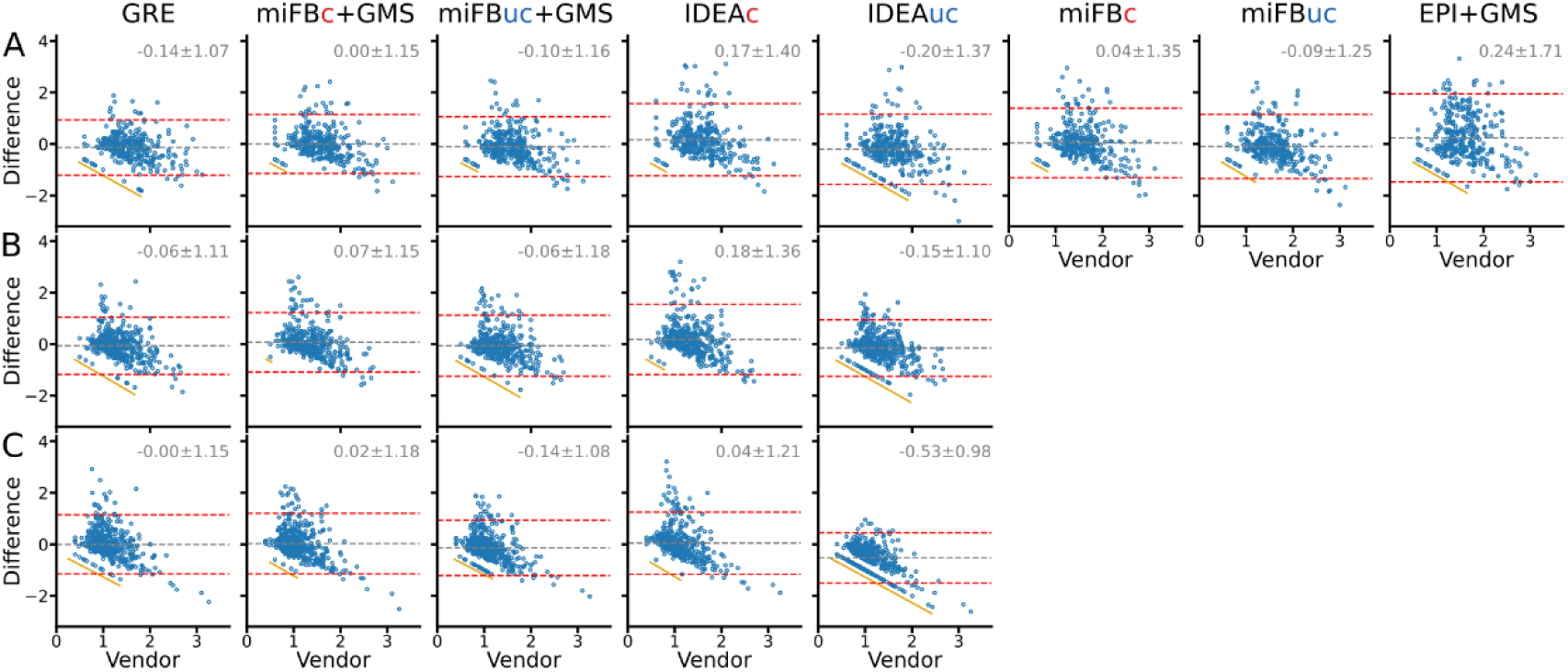
Gold-standard-to-difference plots for MCA diameter in mm for 0.6 mm (A), 0.5 mm (B) and 0.4 mm (C), with Vendor considered as the gold standard. The mean and standard deviation of the differences are shown as dashed lines and printed above. Flow in MCA is predominantly along the frequency encode direction. A linear dependency of the difference to Vendor as indicated by a yellow line originates from segments where vessel information is fully destroyed by flow artifacts, resulting in an estimated vessel diameter of zero.

However, at 0.4 mm resolution (C), differences increased for both uncompensated flyback images (miFBuc, IDEAuc), with a notable increase in the number of missing vessel pixels.

Figure 9 shows gold-standard-to-difference plots for ACA diameter (A) and cross-section area (B), respectively, for GRE, miFB+GMS, and miFB at 0.6 mm resolution. All compared sequences deviated strongly from Vendor in terms of median diameter and cross-section area, with similar distributions observed for GRE and miFB(c,uc)+GMS. Compensated and uncompensated miFB without GMS shows larger deviations from Vendor.

**Figure 9:**
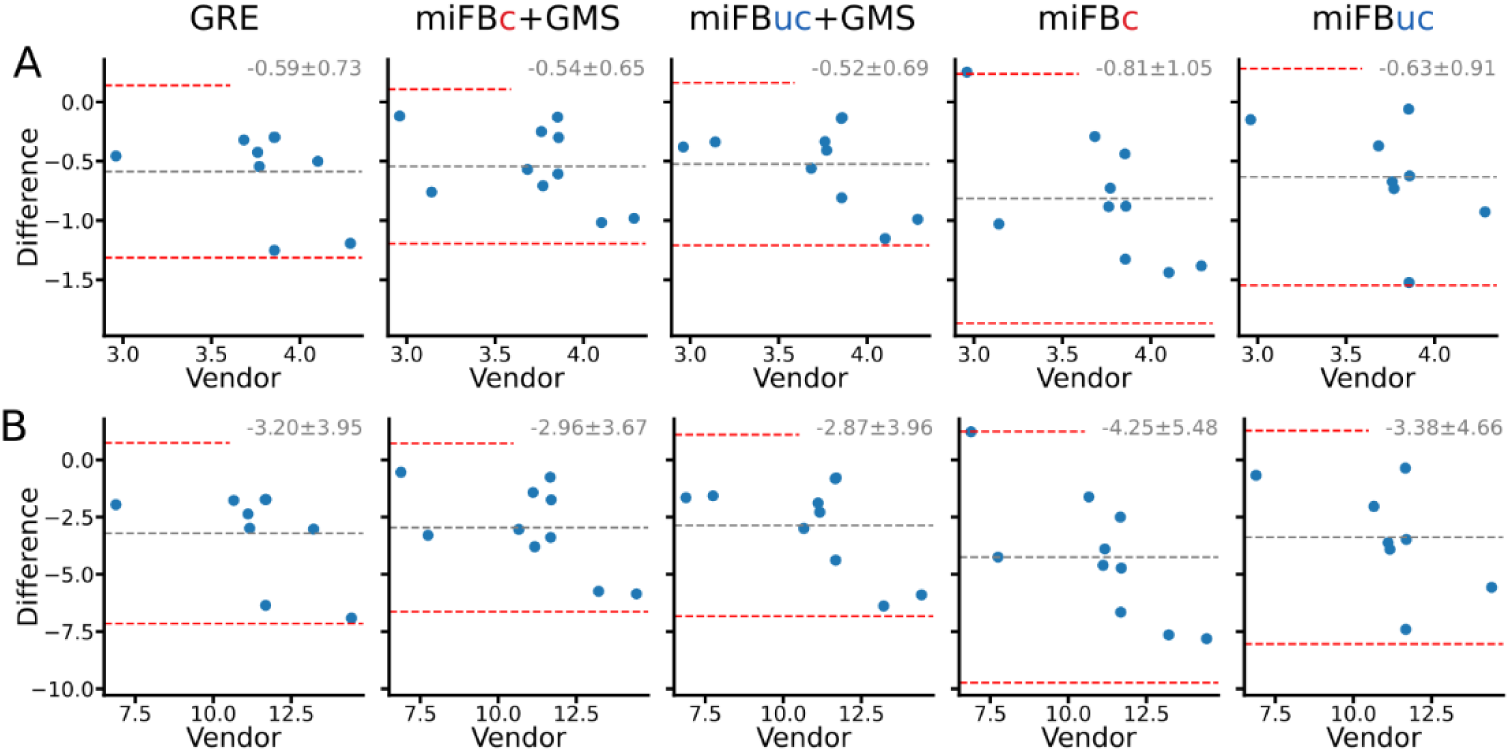
Gold-standard-to-difference plots for ACA diameter in mm (A) and cross-section area in cm2 (B), respectively, for GRE, miFB+GMS, and miFB at 0.6 mm resolution. Vendor is considered as the gold standard. Flow in ACA is predominantly along the phase encode direction. The mean and standard deviation of the differences are shown as dashed lines and printed above. GMS effectively improves the precision of ACA diameter and cross- section area estimation.

## 4. Discussion

Our simulations reveal that for 𝑘_𝑥_ ≠ 0, phase discontinuities related to flow along x persist in both miFB k-spaces, albeit at different positions and frequencies along 𝑘_𝑦_compared to regular EPI (i.e. one peripheral M_1,x_ jump for k_comp_ and two peripheral M_1,x_ jumps for k_uncomp_ with EF=4 in Fig. 1B). The reason for this is that the unsampled echo increases M_1,x_ of the first sampled echo in k_repeated_ the same way as the first echo increases M_1,x_ of the second echo in k_initial_. With miFB, peripheral M_1,x_ jumps generally occur at [2/EF, 4/EF, …] of all lines in k_comp_ and at [1/EF, 3/EF, 5/EF, …] of all lines in k_uncomp_. This difference in the peripheral M_1,x_ evolution, in particular the different k_y_ locations of related phase jumps and the finite M_1,x_ for k_x_=0 may contribute to the increased flow-induced artifacts observed for miFBuc compared to miFBc and should be taken into account for protocol optimization (e.g. choice of phase partial Fourier and EF).

In comparison to EPI+GMS, miFB+GMS significantly reduced flow artifacts from flow along the x-axis, particularly ghosting and signal loss, which can be observed in vessels such as the MCA and PCA. For vessels without flow along the x-axis, we found similar artifact-free results for both EPI+GMS and miFB+GMS. These results were consistently observed in both phantom measurements and images from an exemplary volunteer. Furthermore, when comparing the mean vessel diameter to the vendor-supplied sequence, we observed larger differences for EPI+GMS than miFB+GMS, which supports our other findings. These results match the predictions of our simulations. However, it is essential to consider the trade-offs involved in using the miFB+GMS approach. Repeating each k-space line twice increases the acquisition time by a factor of approximately two. Compared to a regular GRE sequence, the theoretical reduction in acquisition time is not proportional to EF but rather proportional to EF/2. To eliminate sensitivity to discontinuities from alternating readouts, an additional precursor gradient lobe must be added before the first readouts of one of the two interleaves. This ensures that the same moments are obtained in the k-space center for all readouts of one polarity. Without this additional precursor gradient lobe, the resulting flow artifacts would be like those observed with regular EPI. The addition of this gradient increases the TE for miFB by one echo spacing (ESP) compared to regular EPI, which in turn increases acquisition time. Nevertheless, the extra time could be utilized for gradients on other axes, such as GMS gradients along the y-axis or velocity compensation gradients along the z-axis.

The IDEA approach was originally introduced for correction of ghosting from gradient imperfections and off-resonance effects caused by inconsistencies between odd and even echoes. Similar to our method, it also splits odd and even echoes into two k-spaces. In our experiments, we observed slightly worse results for IDEA than for miFB at a resolution of 0.6mm. Although we did not measure IDEA with the GMS approach, with increasing resolution, the major benefit of IDEA becomes apparent: only half of the y-blips are applied, albeit with doubled zeroth-order moment. Since the amplitude of our y-gradient blips is already close to the hardware limits, this increases the duration of the y-blips by a factor of approximately 2 and, as a result, also their first-order moment. The more important consequence, however, is that the number of blips before the k-space center is cut in half, leading to reduced flow artifacts in the images. In our specific case with an EF of 4, only one blip is applied with IDEA. Using a partial Fourier factor of 6/8, this means that no PE-blips are applied before the k-space center is sampled. This leads to significantly reduced artifacts from flow along the y-axis for IDEA compared to miFB. A disadvantage of the approach is that the TE of the two k-spaces and images differs by one ESP.

At a resolution of 0.6mm, miFB+GMS yields similar results as the vendor-supplied TOF sequence and the vendor-based GRE reference sequence for the circle-of-willis. In contrast, Vendor performs significantly better for small vessels outside of the circle-of-willis. A possible reason for this is the dedicated TONE excitation pulse. In addition, velocity compensation is applied along all axes in the vendor-supplied sequence, leading to a reduction in flow artifacts for z-axis flow, as seen in the ICA. A notable reduction in acquisition time is observed with miFB+GMS, which is approximately 2.5 times faster than Vendor. Although to a large degree this reduction is due to the increased Vendor-TR resulting from SAR restrictions, possibly caused by the different excitation schemes.

Similar results were obtained when comparing GRE to miFB+GMS. However, we observed reduced signal at all resolutions for vessels close to the boundaries of the z-FoV. This may be due to the smaller time-bandwidth product of the GRE excitation pulse and imperfect excitation profile near the edge of the z-FoV. Additionally, less slice oversampling was used with GRE, which could increase wrap-around artifacts. Compared to GRE, miFB+GMS achieves a reduction in acquisition time by a factor of about 1.6. The difference between the actual and theoretical reductions of EF/2 can be explained by the increased slice oversampling and the longer duration of parallel imaging reference scans.

Our results confirm the findings of the original GMS paper by Slavin and Riederer (16), which demonstrated a reduction in ghosting artifacts in images acquired with GMS. We observed similar reductions in both phantom and in vivo measurements although we omitted nulling M_1,y_ in the k-space. Notably, ghosting was only evident in vessels oriented along both the x and y axes, but not in those solely aligned with the y direction. This is because flow-induced ghosting propagates along the y-axis independently of vessel angulation and overlaps with signal from vessels along the y-axis. We found that the TE increase due to smoothing gradients with additional velocity compensation in the k-space center can be substantial, particularly at high resolutions and longer echo spacing.

Although the additional time introduced by the precursor gradient lobe on the x-axis could be utilized for gradients on the y-axis, our 0.6mm protocol showed an additional increase in TE of approximately one ESP, if smoothing gradients with flow compensation in the k-space center were applied. This TE increase would result in larger flow artifacts along the readout direction. The trade-off between compensation along the y-axis and reduction of TE requires further evaluation. It is worth noting that it is beneficial to sample the k-space center with shots associated with small ETS, resulting in a smaller M_1_ of the y-blips. This can be accomplished by carefully adjusting the actual phase partial Fourier and segmentation factors.

The alternating polarities of EPI readout gradients induce a phase difference between odd and even echoes at k_x_=0, which can be quantified using formula 3.2. This phase shift, although problematic for regular EPI, provides the additional benefit of velocity encoding for miFB, as it is directly proportional to flow velocity along x. In contrast to IDEA, our miFB approach allows the exploitation of this inherent velocity encoding of the readout gradients in EPI: since both images reconstructed from k_comp_ and k_uncomp_ share the same TE, it enables the quantification of velocity by subtracting the reconstructed miFBuc and miFBc phase images (36). An example can be found in Figure S1.

### Limitations

As resolution increases, artifacts become more pronounced in uncompensated images, whereas compensated images remain relatively artifact-free for flow along the x-axis. This becomes particularly evident at 0.4 mm resolution, where flow along the x-axis induced artifacts are significantly larger for the uncompensated compared to the compensated case. Computing the mean across both images can lead to a loss of accuracy in the resulting mean image. This limitation reduces the scanning efficiency by a factor of two, as it hinders the use of both flyback images. However, this issue only arises at very high resolutions, where the phase offset in the center of the uncompensated k-space and peripheral phase jumps become substantial enough to cause significant flow-induced artifacts. With increasing voxel size, differences between the compensated and mean images diminish and become negligible at 0.6 mm and 0.8 mm in-plane isotropic resolution (Figure 7 B).

At high resolutions, large flow artifacts persist in particular for flow along the y-axis. Compensating related first order moments at the k-space center could potentially reduce these artifacts. Furthermore, all EPI-based and GRE images exhibited artifacts from flow along the z-direction. Implementing flow compensation along z, taking advantage of the available EPI precursor/ESP dead-time, would likely improve the results, as it would help to minimize the effects of flow on image quality.

## 5. Conclusion

In conclusion, our proposed miFB sampling has demonstrated significant reductions in flow-induced artifacts, particularly ghosting and signal loss, compared to conventional EPI. The combination of miFB with GMS further enhances its performance, yielding comparable results to vendor-supplied sequences at a resolution of 0.6mm. Although our method is effective at reducing artifacts, it is worth noting that signal loss in uncompensated images can become a limiting factor for scanning efficiency at very high resolutions (e.g., smaller voxels than 0.6 mm in-plane isotropic). Additionally, our results suggest that compensating or reducing first-order moments along the y- and z-directions may be necessary to further reduce flow-induced artifacts.

## Acknowledgments

### Author contributions

The authors thank Pål-Erik Goa for valuable discussions. This work received financial support from Helmholtz Association Initiative and Networking Fund, funding code ZT-IPF- 4-042 (Helmholtz Imaging Project “HighLine”).

### Financial disclosure

None reported.

### Data Availability Statement

In support of Magnetic Resonance in Medicine’s reproducible research goal, the Python-based simulation code has been made available at https://github.com/mrphysics-bonn/miFB-simulations.

## Supporting information

The following supporting information is available as part of the online article:

## Bibliography

1. Sakata A, Fushimi Y, Okada T, et al. Evaluation of cerebral arteriovenous shunts: a comparison of parallel imaging time-of-flight magnetic resonance angiography (TOF-MRA) and compressed sensing TOF-MRA to digital subtraction angiography. Neuroradiology. 2021;63(6):879–887. doi:10.1007/s00234-020-02581-y

2. Lin Z, Zhang X, Guo L, et al. Clinical feasibility study of 3D intracranial magnetic resonance angiography using compressed sensing. Journal of Magnetic Resonance Imaging. 2019;50(6):1843–1851. doi:10.1002/jmri.26752

3. Firmin DN, Klipstein RH, Hounsfield GL, Paley MP, Longmore DB. Echo-planar high-resolution flow velocity mapping. Magnetic Resonance in Med. 1989;12(3):316–327. doi:10.1002/mrm.1910120304

4. Wielopolski PA, Manning WJ, Edelman RR. Single breath-hold volumetric imaging of the heart using magnetization-prepared 3-dimensional segmented echo planar imaging. Journal of Magnetic Resonance Imaging. 1995;5(4):403–409. doi:10.1002/jmri.1880050406

5. Slavin GS, Riederer SJ, Ehman RL. Two-dimensional multishot echo-planar coronary MR angiography. Magnetic Resonance in Medicine. 1998;40(6):883–889. doi:10.1002/mrm.1910400614

6. Breuer F a, Blaimer M, Heidemann RM, Mueller MF, Griswold M a, Jakob PM. Controlled aliasing in parallel imaging results in higher acceleration (CAIPIRINHA) for multi-slice imaging. Magnetic resonance in medicine : official journal of the Society of Magnetic Resonance in Medicine / Society of Magnetic Resonance in Medicine. 2005;53(3):684–691. doi:10.1002/mrm.20401

7. Setsompop K, Gagoski BA, Polimeni JR, Witzel T, Wedeen VJ, Wald LL. Blipped-controlled aliasing in parallel imaging for simultaneous multislice echo planar imaging with reduced g-factor penalty. Magnetic Resonance in Medicine. 2012;67(5):1210–1224. doi:10.1002/mrm.23097

8. Hendriks AD, D’Agata F, Raimondo L, et al. Pushing functional MRI spatial and temporal resolution further: High-density receive arrays combined with shot-selective 2D CAIPIRINHA for 3D echo-planar imaging at 7 T. NMR in Biomedicine. 2020;33(5):e4281. doi:10.1002/nbm.4281

9. Norbeck O, Sprenger T, Avventi E, et al. Optimizing 3D EPI for rapid T1 -weighted imaging. Magn Reson Med. 2020;84(3):1441–1455. doi:10.1002/mrm.28222

10. Stirnberg R, Stöcker T. Segmented K-space blipped-controlled aliasing in parallel imaging for high spatiotemporal resolution EPI. Magnetic Resonance in Med. 2021;85(3):1540–1551. doi:10.1002/mrm.28486

11. Duerk JL, Simonetti OP. Theoretical aspects of motion sensitivity and compensation in echo-planar imaging. Magnetic Resonance Imaging. 1991;1(6):643–650. doi:10.1002/jmri.1880010605

12. Butts K, Riederer SJ. Analysis of flow effects in echo-planar imaging. Magnetic Resonance Imaging. 1992;2(3):285–293. doi:10.1002/jmri.1880020306

13. Nishimura DG, Jackson JI, Pauly JM. On the nature and reduction of the displacement artifact in flow images. Magnetic Resonance in Med. 1991;22(2):481–492. doi:10.1002/mrm.1910220255

14. Nishimura DG, Irarrazabal P, Meyer CH. A velocityk-space analysis of flow effects in echo-planar and spiral imaging. Magn Reson Med. 1995;33(4):549–556. doi:10.1002/mrm.1910330414

15. Pat GTL, Meyer CH, Pauly JM, Nishimura DG. Reducing flow artifacts in echo-planar imaging. Magnetic Resonance in Med. 1997;37(3):436–447. doi:10.1002/mrm.1910370323

16. Slavin GS, Riederer SJ. Gradient moment smoothing: A new flow compensation technique for multi-shot echo-planar imaging. Magnetic Resonance in Med. 1997;38(3):368–377. doi:10.1002/mrm.1910380304

17. Beck G, Li D, Haacke EM, Noll TG, Schad LR. Reducing oblique flow effects in interleaved EPI with a centric reordering technique. Magnetic Resonance in Med. 2001;45(4):623–629. doi:10.1002/mrm.1084

18. Jaffer FA, Wen H, Jezzard P, Balaban RS, Wolff SD. Centric ordering is superior to gradient moment nulling for motion artifact reduction in EPI. Journal of Magnetic Resonance Imaging. 1997;7(6):1122–1131. doi:10.1002/jmri.1880070627

19. Blömer S, Stirnberg R, Stöcker T. Highly segmented skipped-CAIPI 3D-EPI with a modified interleaved Flyback and partial Fourier acquisition for fast MR angiography. In: Proceedings of the 2024 ISMRM & SMRT Annual Meeting & Exhibition. ; 2024.

20. Poser BA, Barth M, Goa P, Deng W, Stenger VA. Single-shot echo-planar imaging with Nyquist ghost compensation: Interleaved dual echo with acceleration (IDEA) echo-planar imaging (EPI). Magnetic Resonance in Med. 2013;69(1):37–47. doi:10.1002/mrm.24222

21. Feinberg DA, Oshio K. Phase errors in multi-shot echo planar imaging. Magnetic Resonance in Med. 1994;32(4):535–539. doi:10.1002/mrm.1910320418

22. Stirnberg R, Deistung A, Reichenbach JR, Breteler MMB, Stöcker T. Rapid submillimeter QSM and R_2_* mapping using interleaved multishot 3D-EPI at 7 and 3 Tesla. Magnetic Resonance in Med. Published online July 10, 2024. doi:10.1002/mrm.30216

23. Farzaneh F, Riederer SJ. Hybrid imaging with gradient recalled sliding echoes. In: Proc., SMRI, 7th Annual Meeting, 1989. ; 1989:70.

24. Marques JP, Kober T, Krueger G, Van Der Zwaag W, Van De Moortele PF, Gruetter R. MP2RAGE, a self bias-field corrected sequence for improved segmentation and T1- mapping at high field. NeuroImage. 2010;49(2):1271–1281. doi:10.1016/j.neuroimage.2009.10.002

25. Griswold MA, Jakob PM, Heidemann RM, et al. Generalized autocalibrating partially parallel acquisitions (GRAPPA). Magnetic Resonance in Med. 2002;47(6):1202–1210. doi:10.1002/mrm.10171

26. Breuer FA, Blaimer M, Mueller MF, et al. Controlled aliasing in volumetric parallel imaging (2D CAIPIRINHA). Magnetic Resonance in Med. 2006;55(3):549–556. doi:10.1002/mrm.20787

27. Tustison NJ, Avants BB, Cook PA, et al. N4ITK: Improved N3 Bias Correction. IEEE Trans Med Imaging. 2010;29(6):1310–1320. doi:10.1109/TMI.2010.2046908

28. Manjón JV, Coupé P, Martí-Bonmatí L, Collins DL, Robles M. Adaptive non-local means denoising of MR images with spatially varying noise levels. Magnetic Resonance Imaging. 2010;31(1):192–203. doi:10.1002/jmri.22003

29. Tustison NJ, Cook PA, Holbrook AJ, et al. The ANTsX ecosystem for quantitative biological and medical imaging. Sci Rep. 2021;11(1):9068. doi:10.1038/s41598-021-87564-6

30. Avadiappan S, Payabvash S, Morrison MA, Jakary A, Hess CP, Lupo JM. A Fully Automated Method for Segmenting Arteries and Quantifying Vessel Radii on Magnetic Resonance Angiography Images of Varying Projection Thickness. Front Neurosci. 2020;14:537. doi:10.3389/fnins.2020.00537

31. vmtk/vmtk. Published online May 30, 2025. Accessed June 5, 2025. https://github.com/vmtk/vmtk

32. 3D Slicer image computing platform. 3D Slicer. June 5, 2025. Accessed June 5, 2025. https://slicer.org/

33. Fedorov A, Beichel R, Kalpathy-Cramer J, et al. 3D Slicer as an Image Computing Platform for the Quantitative Imaging Network. Magn Reson Imaging. 2012;30(9):1323–1341. doi:10.1016/j.mri.2012.05.001

34. Kikinis R, Pieper SD, Vosburgh KG. 3D Slicer: A Platform for Subject-Specific Image Analysis, Visualization, and Clinical Support. In: Jolesz FA, ed. Intraoperative Imaging and Image-Guided Therapy. Springer; 2014:277–289. doi:10.1007/978-1-4614-7657-3_19

35. Kapur T, Pieper S, Fedorov A, et al. Increasing the impact of medical image computing using community-based open-access hackathons: The NA-MIC and 3D Slicer experience. Medical Image Analysis. 2016;33:176–180. doi:10.1016/j.media.2016.06.035

36. Blömer S, Stirnberg R, Stöcker T. Inherent velocity encoding in EPI-based phase contrast MRI using a modified flyback approach. In: Proceedings of the 2025 ISMRM & SMRT Annual Meeting & Exhibition. ; 2025.

